# Levodopa Preferentially Reorganizes Dorsal Subthalamic Activity in Parkinson’s Disease

**DOI:** 10.64898/2026.10.13.26365259

**Authors:** Sandeep Sathyanandan Nair, Prajakta Joshi, Veronika Filyushkina, Ksenia Sayfulina, Aratrik Guha, Alexey Sedov, Aasef G. Shaikh

**Affiliations:** Department of Biomedical Engineering, Case Western Reserve University, Cleveland, OH 44106, USA; Cleveland FES Center, Louis Stokes Cleveland VA Medical Center, Cleveland, OH 44106, USA; N.N. Semenov Federal Research Center for Chemical Physics Russian Academy of Sciences, Moscow, Russia; Moscow Institute of Physics and Technology, Dolgoprudny, Moscow Region, Russia; Department of Neurology, University Hospitals, Cleveland, OH 44106, USA; Department of Neurology, Case Western Reserve University, Cleveland, OH 44106, USA

**Keywords:** Parkinson’s disease, subthalamic nucleus, beta oscillations, local field potentials, dopaminergic medication, deep brain stimulation, spatial organization

## Abstract

**Background:** Dopamine replacement reshapes subthalamic activity in Parkinson’s disease, but whether this response reflects anatomically organized network reconfiguration remains unclear.

**Objectives:** To determine whether levodopa preferentially reorganizes dorsal subthalamic spectral activity and cortico-subthalamic directionality.

**Methods:** Twelve participants undergoing bilateral subthalamic deep brain stimulation contributed 30 contacts (14 dorsal, 16 ventral). Recordings obtained OFF and ON medication underwent spectral parameterization and multiband state-space analysis. Simultaneous electroencephalography in six participants enabled assessment of directional cortico-subthalamic coupling using the phase slope index.

**Results:** Levodopa suppressed beta power more strongly dorsally than ventrally (−2.473 versus −0.120 dB; p=0.0438). Dorsal theta power increased, with a trend toward an increase after baseline normalization (p=0.07). Dorsal contacts exhibited greater theta–beta (+5.904 versus +1.610 dB; p=0.006) and gamma–beta (+2.516 versus +0.384 dB; p=0.0188) state-space shifts, principally driven by beta suppression. Gamma changes did not differ regionally. Concurrently, dorsal low-beta and beta interactions shifted toward cortical leading at selected electrodes, whereas ventral responses were less consistent.

**Conclusions:** Levodopa produces a topographically organized subthalamic response, combining preferential dorsal beta suppression with altered cortico-subthalamic directionality. These findings position contact anatomy as a key consideration in characterizing dopaminergic network responses and motivate clinical validation of spatially informed multiband biomarkers for adaptive stimulation.

## Introduction

Parkinson’s disease (PD), first described as the “shaking palsy”^1^, was initially explained by impaired motor activation, with dopamine loss thought to increase basal-ganglia output and reduced thalamocortical drive^2-5^. This framework later expanded to include altered firing patterns, bursting, oscillations, and synchronization across basal-ganglia circuits^6-9^. Human neurophysiological recordings further showed dopamine-dependent synchronization within basal-ganglia and cortico-subthalamic networks^10-12^, increasingly characterizing PD as a distributed network disorder whose pathophysiology extends beyond the basal ganglia to engage cortical, thalamic, brainstem, and cerebellar systems^13-17^.

Animal studies showed dopaminergic modulation of parkinsonian physiology across firing rate, regularity, oscillatory state, and synchronization, but not uniformly across nuclei^2, 18-20^. Human subthalamic nucleus (STN) local field potential (LFP) recordings confirmed exaggerated low-frequency synchronization, with levodopa suppressing activity below 30 Hz and promoting higher-frequency activity^10^. Simultaneous recordings also identified 14–30Hz (beta-band) STN synchronization attenuated by movement and dopaminergic therapy^11^. Beta-band activity was particularly prominent in the dorsal STN, where oscillatory neuronal discharge was strongly coherent with the LFP ^21^. Importantly, levodopa-induced suppression of 8–35-Hz STN activity correlated with improvement in contralateral bradykinesia and rigidity, directly linking the physiological response to clinical benefit^22^. Later work showed that prolonged beta bursts track worse motor impairment and are shortened by levodopa^23^. State-based spectral dynamics predicted impairment better than beta power alone^24^.

These abnormalities are spatially organized within the STN, with beta activity concentrated in dorsal sensorimotor regions while ventral STN shows relatively greater gamma activity^21, 25, 26^. Levodopa also modulates intranuclear communication, altering functional connectivity and ventral-to-dorsal information flow within the STN in association with motor improvement^27^. More detailed mapping supports frequency-specific dorso-ventral segregation within the STN^21, 25, 26, 28^. These data support a topographically organized model of dopamine-sensitive STN physiology^27, 29^. Direct evidence for such differentiation, however, remains limited. We hypothesized that levodopa produces stronger prokinetic spectral modulation and cortex-to-STN directional drive in dorsal compared to ventral STN. We tested the hypothesis using spatially resolved LFPs, spectral parameterization, state-space analysis, and directional connectivity.

## Methods

### Participants and recordings

Twelve individuals with PD undergoing bilateral STN DBS participated. The study was approved by institutional ethics committees at Louis Stokes Cleveland VA Medical Center, and Burdenko Neurosurgical Institute. The participants signed informed consent form prior to participating in this study. Demographic data are shown in Table 1. LFPs were recorded from available DBS contacts in medication-OFF and medication-ON states. Six participants underwent acute postoperative recordings using an externalized system. Scalp electroecephalography (EEG) was measured from these participants. Remaining six underwent chronic recordings with a sensing-enabled implanted pulse generator. Data from both hemispheres were included when of sufficient quality.

**Table 1.** Demographic and clinical characteristics of patients with Parkinson’s disease. Patient characteristics include sex, age, levodopa equivalent daily dose (LEDD), and Unified Parkinson’s Disease Rating Scale Part III (UPDRS-III) scores in the OFF and ON medication states. NA, not available.

| Patient ID | Gender | Age | LEDD | UPDRS-III<br>OFF | UPDRS-III<br>ON |
| --- | --- | --- | --- | --- | --- |
| PD01 | F | 51-55 | 1350 mg | 32 | 18 |
| PD02 | M | 36-40 | 2250 mg | 40 | 16 |
| PD03 | M | 51-55 | 1820 mg | 47 | 14 |
| PD04 | F | 46-50 | 2350 mg | 36 | 14 |
| PD05 | M | 61-65 | 1857 mg | 41 | 35 |
| PD06 | M | 46-50 | 1875mg | 40 | 16 |
| PD07 | M | 56-60 | 1505 mg | 42 | 18 |
| PD08 | M | 76-80 | 635 mg | 40 | 19 |
| PD09 | M | 76-80 | 1000 mg | 45 | 21 |
| PD10 | F | 56-60 | 600 mg | 38 | 18 |
| PD11 | M | 66-70 | 400 mg | 28 | 12 |
| PD12 | M | 51-55 | 1200 mg | 46 | 17 |

### Electrode localization

Electrode locations were reconstructed by co-registering postoperative computed tomography (CT) with preoperative magnetic resonance imaging (MRI). Trajectories and contacts were localized within patient-specific STN anatomy using Lead-DBS^29-31^. For primary analyses, contacts were classified as dorsal or ventral relative to the STN anatomical midpoint.

### Spectral analysis

LFPs were analyzed separately in medication-OFF and medication-ON states. Power spectral density (PSD) was estimated for each recording and parameterized with FOOOF^32^ to separate aperiodic and oscillatory components. Power was quantified in delta (0.5-4Hz), theta (4-8Hz), alpha (8-13Hz), beta (14-30Hz), and gamma (>30Hz) frequency bands. Medication effects were calculated as medication-ON minus medication-OFF for each contact, and frequency-resolved analyses characterized changes across the spectrum.

### Neural state-space analysis

To assess whether medication effects differed by STN subregion, theta–beta and gamma–beta changes were represented in two-dimensional state spaces. For each contact, medication-related changes were defined relative to baseline as (Δθ, Δβ) and (Δγ, Δβ), where positive Δθ or Δγ indicates increased theta or gamma power and negative Δβ indicates beta suppression. Thus, movement toward the positive-theta/negative-beta or positive-gamma/negative-beta quadrant represented the hypothesized therapeutic direction. Group distributions were visualized with individual data points, kernel-density contours, covariance ellipses, and means. The primary summary metric was projection along the therapeutic axis: *P*_*θβ*_ = Δ*θ* − Δ*β* and *P*_*γβ*_ = Δ*γ* − Δ*β*, such that higher values indicate greater theta/gamma increase and/or beta suppression. Dorsal and ventral contacts were compared at the contact level using two-sided Wilcoxon rank-sum tests for Δθ, Δβ, and Pθβ, and separately for Δγ, Δβ, and Pγβ.

### Statistical analysis

Analyses combined both recording cohorts, with contacts grouped by dorsoventral STN location. Medication-related spectral changes were assessed within each contact by comparing medication-OFF and medication-ON recordings. For each frequency band, OFF versus ON power was compared using Wilcoxon signed-rank tests. Tests were two-sided for delta, theta, alpha, and gamma bands; for beta, a one-sided Wilcoxon signed-rank test was used based on the prespecified hypothesis of beta suppression with levodopa. Significance was defined as p<0.05. Frequency-resolved effects were also quantified as ΔPSD=medication-ON−baseline and summarized separately for dorsal and ventral contacts. The combined cohort included 14 dorsal and 16 ventral contacts.

## Results

The results tested our hypothesis that dopaminergic therapy would produce greater state-space modulation of STN spectral activity favoring prokinetic effects and stronger cortex-to-STN directional drive in dorsal than ventral STN. Figure 1 illustrates electrode localization into dorsal versus ventral with anatomical reference (Figure 1A) and medication-related spectral changes in a representative PD participant (Figure 1B1-B6). Fitting oscillations and one-over-f (FOOOF) parameterization separated the oscillatory component from the aperiodic background, allowing medication-related changes in frequency-specific oscillatory activity to be examined independently of the broadband spectral component. In the representative dorsal contact, medication-on was associated with reduced beta-band and increased theta-band oscillatory power compared with off medication state (Figure 1B2). In addition, there was an offset in aperiodic component in response to dopaminergic medication (Figure 1B3). Such periodic spectral changes were not seen in ventral STN recordings (Figure 1B5); however, aperiodic offset was still noteworthy in the ventral STN contact (Figure 1B6).

**Figure 1.**
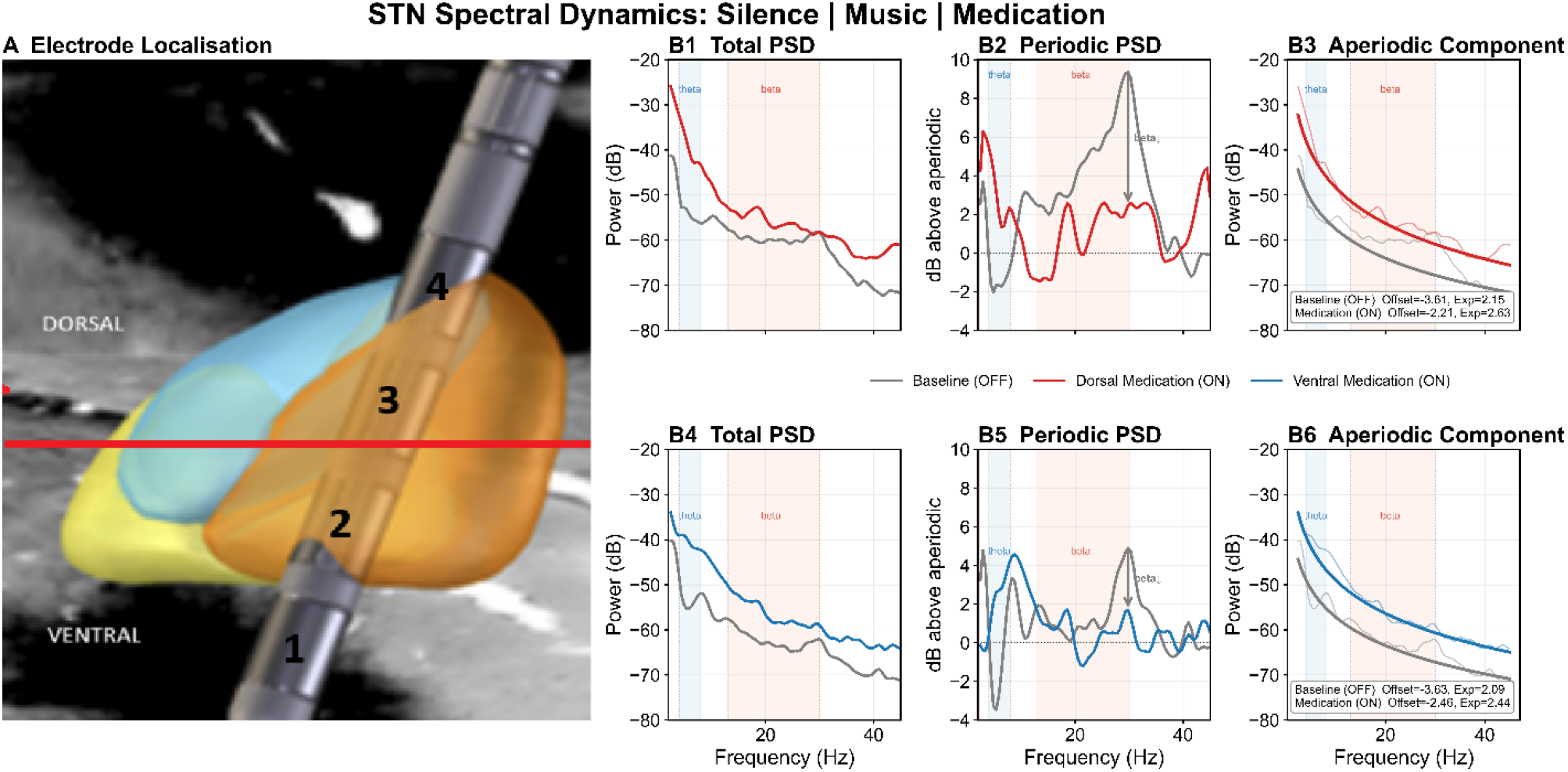
Representative electrode localization and medication-related changes in STN LFP spectra. (A) Three-dimensional reconstruction of the bilateral DBS electrodes relative to the STN and surrounding anatomical structures in a representative participant. The red horizontal line indicates the anatomical midpoint used to define the dorsoventral classification of recording contacts. (B) Representative spectral analysis from two recording contacts in the same participant. B1,B2,B3 represent recordings from dorsal contacts, while the B4,B5,B6 represent the recordings from the ventral contacts. All recordings are from the electrode in the Right STN. Left, raw power spectral density (PSD) during medication-off (blue, dashed) and medication-on (red) conditions. Middle, FOOOF-derived oscillatory power after separation of the aperiodic component, demonstrating medication-related changes in frequency-specific oscillatory activity. Right, fitted aperiodic components for medication-off and medication-on recordings, with corresponding aperiodic parameters indicated. Together, the panels illustrate the anatomical localization and spectral parameterization used to quantify medication-related changes in STN activity.

The exemplary changes noted in Figure 1 were consistently seen across the cohort of 30 STN contacts (14 dorsal and 16 ventral) from 24 hemispheres in 12 PD participants. To compare the effects across participants, we defined ΔPSD as dopamine therapeutic minus dopamine depleted state, such that negative values indicate medication-associated suppression and positive values indicate an increase relative to the off medication baseline.

The frequency-resolved analysis revealed a clear anatomical and frequency-dependent pattern of levodopa-related modulation (Figure 2A). Band-specific analysis confirmed significant levodopa-related modulation of dorsal STN activity in both the theta and beta frequency ranges (Figure 2B). Beta showed the strongest effect, with beta power significantly reduced during levodopa ON compared with baseline/OFF (Figure 2B, p<0.001, one-sided Wilcoxon signed-rank test), consistent with robust levodopa-associated beta suppression. A significant increase in theta power was also observed during levodopa ON (Figure 2B, p<0.05, two-sided Wilcoxon signed-rank test). The levodopa-related changes in dorsal STN were insignificant in delta (p=0.391), alpha (p=0.761), or gamma (p=0.426) bands (Figure 2B).

**Figure 2.**
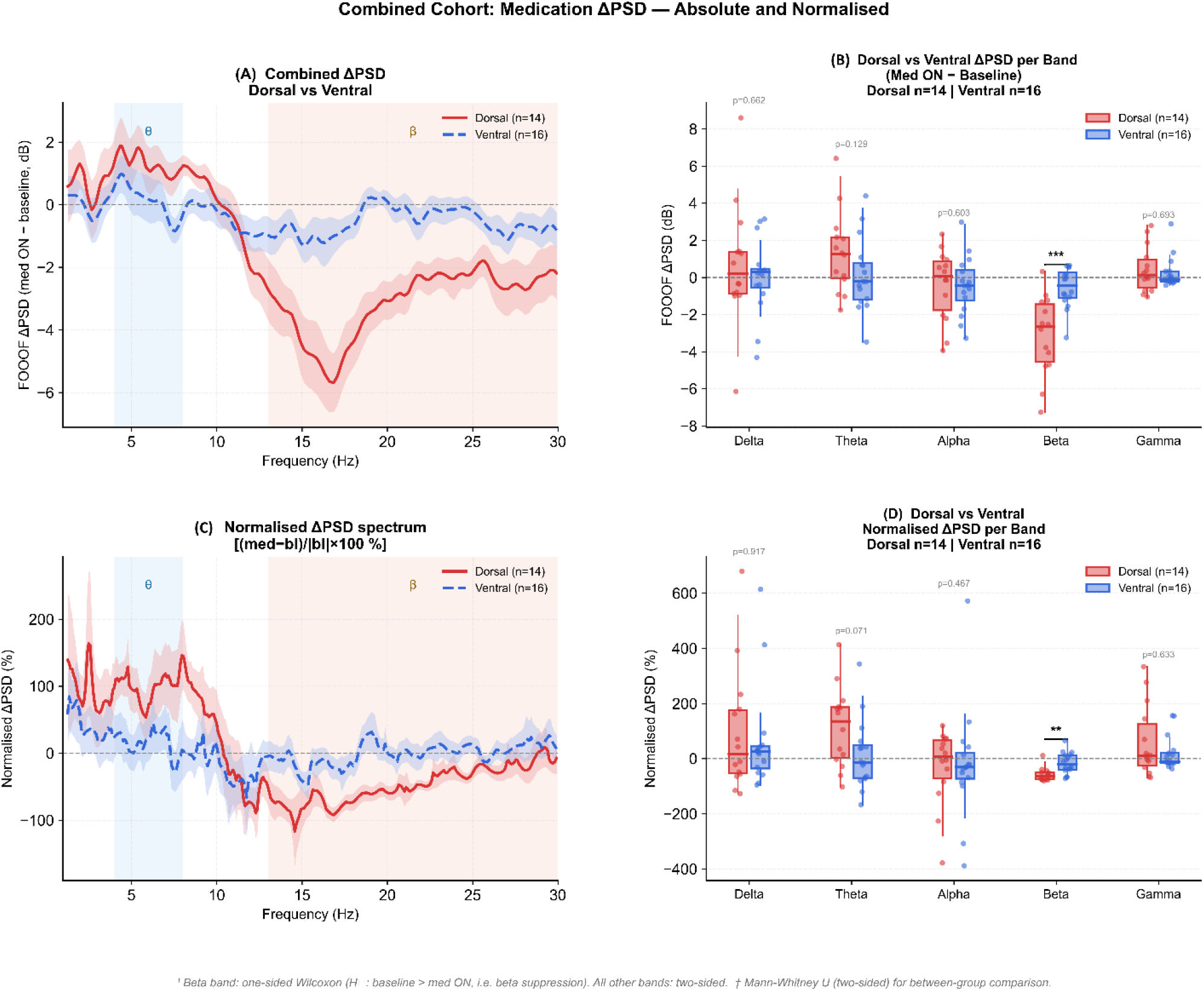
Frequency- and anatomical location-dependent effects of dopaminergic medication on STN spectral activity. (A) Frequency-resolved change in FOOOF-derived spectral power (ΔPSD; medication-ON minus medication-OFF) for dorsal (red) and ventral (blue) STN contacts across the 1–30 Hz frequency range. Shaded regions represent the corresponding variability across contacts. The largest separation between dorsal and ventral responses occurs within the beta range. (B) Medication-related changes in FOOOF-derived power across delta, theta, alpha, beta, and gamma bands in dorsal versus ventral STN contacts. (C) Normalized frequency-resolved changes in FOOF-derived spectral power (normalized ΔPSD; (medication-ON minus medication-OFF)/(medication-OFF)). (D) normalized medication related changes in FOOF-derived power across delta, theta, alpha, beta, and gamma bands in dorsal versus ventral STN contacts. For within-region comparisons, a one-sided Wilcoxon signed-rank test was used for beta (prespecified direction: medication-OFF > medication-ON) and two-sided Wilcoxon signed-rank tests were used for the remaining frequency bands. Negative ΔPSD values indicate medication-induced suppression. *p < 0.05; **p < 0.01; ***p < 0.001.

Spectral power, even at baseline, may be spatially discrete within the STN. To account for baseline heterogeneity in the spectral power on medication effects, we examined whether results observed in Figure 2A,B remains consistent after normalizing to baseline. As depicted in Figure 2C,D the spatially diverse effects of levodopa on STN spectral activity remains even after normalizing the activity to baseline power, with a caveat that dorsal STN theta activity increased but statistical significance with 95% confidence was not met (p = 0.07). The reduction in dorsal STN beta band still remained significant (p<0.01). The ventral STN spectral changes were insignificant (Figure 2C,D).

### Neural state-space analysis: preferential levodopa-induced reorganization in dorsal STN theta-beta state space

Levodopa produced a larger increase in theta power at dorsal than ventral contacts, although the difference did not reach statistical significance (dorsal: +3.431 dB; ventral: +1.491 dB; Wilcoxon rank-sum, p=0.140). In contrast, the reduction in beta power was significantly greater at dorsal contacts (dorsal: −2.473 dB; ventral: −0.120 dB; p=0.0438) (Figure 3A,B).

**Figure 3.**
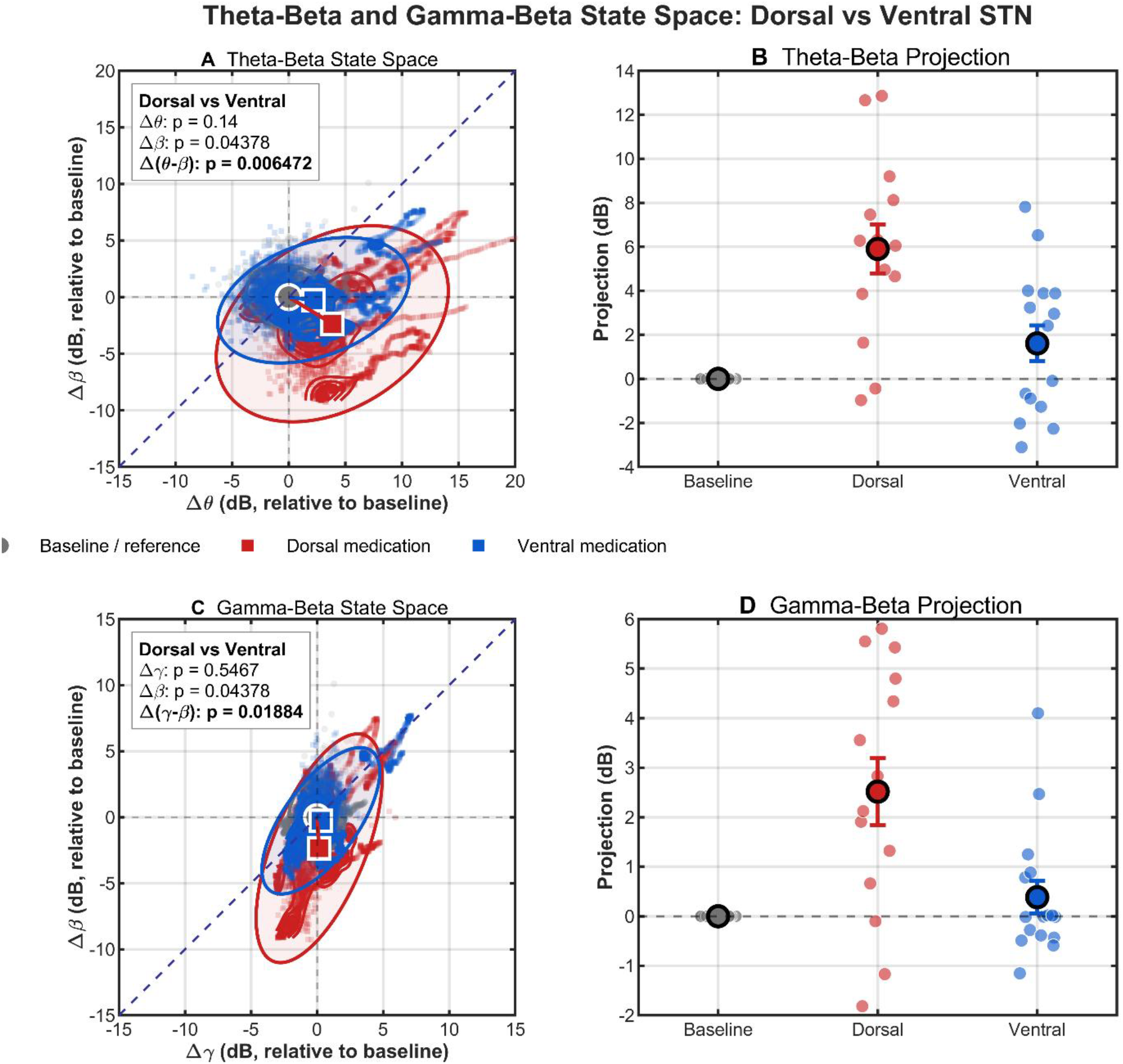
Medication-induced spectral shifts in dorsal and ventral STN contacts. (A) Theta-beta state space showing contact-level baseline and medication distributions for dorsal and ventral STN. The x-axis represents change in theta power, and the y-axis change in beta power relative to baseline. Arrows indicate the transition between mean baseline and medication states. The dashed diagonal indicates equal theta and beta changes. The inset reports contact-level Wilcoxon rank-sum comparisons of medication-induced changes between dorsal and ventral contacts. (B) Theta-beta projection, defined as, with positive values indicating movement toward increased theta and reduced beta. (C–D) Corresponding gamma-beta state space and projection, with the projection defined as. Red indicates dorsal STN contacts and blue indicates ventral STN contacts; gray indicates baseline/reference values. Data combine Cohorts 1 and 2 and are analyzed at the contact level.

When theta and beta changes were considered jointly using the theta-beta projection, dorsal contacts demonstrated a significantly greater levodopa-induced shift toward the hypothesized state than ventral contacts. The mean change in the theta-beta projection was +5.904 dB at dorsal contacts compared with +1.610 dB at ventral contacts (p=0.006).

This pattern indicates that the stronger dorsal shift in theta-beta state space was driven primarily by the substantially greater reduction in beta power, with a numerically larger increase in theta providing an additional contribution.

### Gamma-beta state space

The medication-induced change in gamma power did not differ significantly between dorsal and ventral contacts (dorsal: +0.043 dB; ventral: +0.265 dB; p=0.547) (Figure 3C,D). As in the theta-beta analysis, beta suppression was significantly greater at dorsal contacts than at ventral contacts (dorsal: −2.473 dB; ventral: −0.120 dB; p=0.0438). Despite the absence of a significant dorsal-ventral difference in gamma change, the combined gamma-beta projection showed a significantly greater medication-induced shift at dorsal contacts. The mean change in gamma-beta projection was +2.516 dB for dorsal contacts compared with +0.384 dB for ventral contacts (p=0.0188). Thus, the dorsal predominance in the gamma-beta state-space shift was again primarily associated with stronger beta suppression rather than a differential increase in gamma power.

Taken together, these findings indicate that dorsal STN has a stronger levodopa-induced displacement along the therapeutic axis than ventral STN as summarized in Supplementary Table 1. The effect was most evident when theta/gamma changes and beta suppression were considered jointly rather than as isolated frequency bands. In particular, the significant dorsal-ventral difference in both the theta-beta and gamma-beta projections indicates that the anatomical distinction is captured most clearly by the combined reorganization of oscillatory activity, with beta suppression providing a major contribution to the stronger dorsal shift.

### Directional Cortico-STN Connectivity Analysis

To characterize the directionality of oscillatory interactions between cortical activity and the STN, we estimated the phase slope index (PSI) between EEG and STN LFP recordings. The PSI provides a frequency-resolved measure of directional information flow from the phase relationship between two signals. In the present analysis, PSI was calculated separately between cortical EEG channels and STN LFP recordings from dorsal and ventral STN regions. The analysis was performed separately for theta, alpha, low beta (14–20Hz), beta (14–30Hz), and gamma. For each band and recording site, PSI was calculated for baseline (OFF-levodopa) and therapeutic levodopa conditions to quantify the direction and magnitude of cortico-STN coupling. For interpretation, PSI was expressed using the EEG→STN convention: positive values indicate cortical activity leading STN activity, negative values indicate STN activity leading cortical activity, and values near zero indicate weak or negligible directional asymmetry. This convention was maintained for both dorsal and ventral STN recordings.

#### Comparison Between Medication Conditions

Levodopa-related changes in directional cortico-STN coupling were quantified as the within-subject difference between the levodopa-on and levodopa-off conditions (Δ*PSI*). Accordingly, a positive Δ*PSI* indicates a shift toward greater cortex-to-STN directionality following medication, whereas a negative Δ*PSI* indicates a shift toward greater STN-to-cortex directionality.

#### Dorsal STN

For recordings from the dorsal STN, baseline (levodopa-off) and therapeutic levodopa conditions showed substantial inter-individual variability in PSI across cortical electrodes and frequency bands (Figure 4, top row). At the group level, the dominant directional relationship varied according to frequency and cortical location. Low-beta and beta activity had predominantly positive PSI values at several cortical sites, indicating a tendency toward cortex-leading-STN interactions in these frequency ranges. In contrast, negative PSI values were also observed in individual subjects, demonstrating considerable heterogeneity in the directionality of cortico-STN coupling.

**Figure 4.**
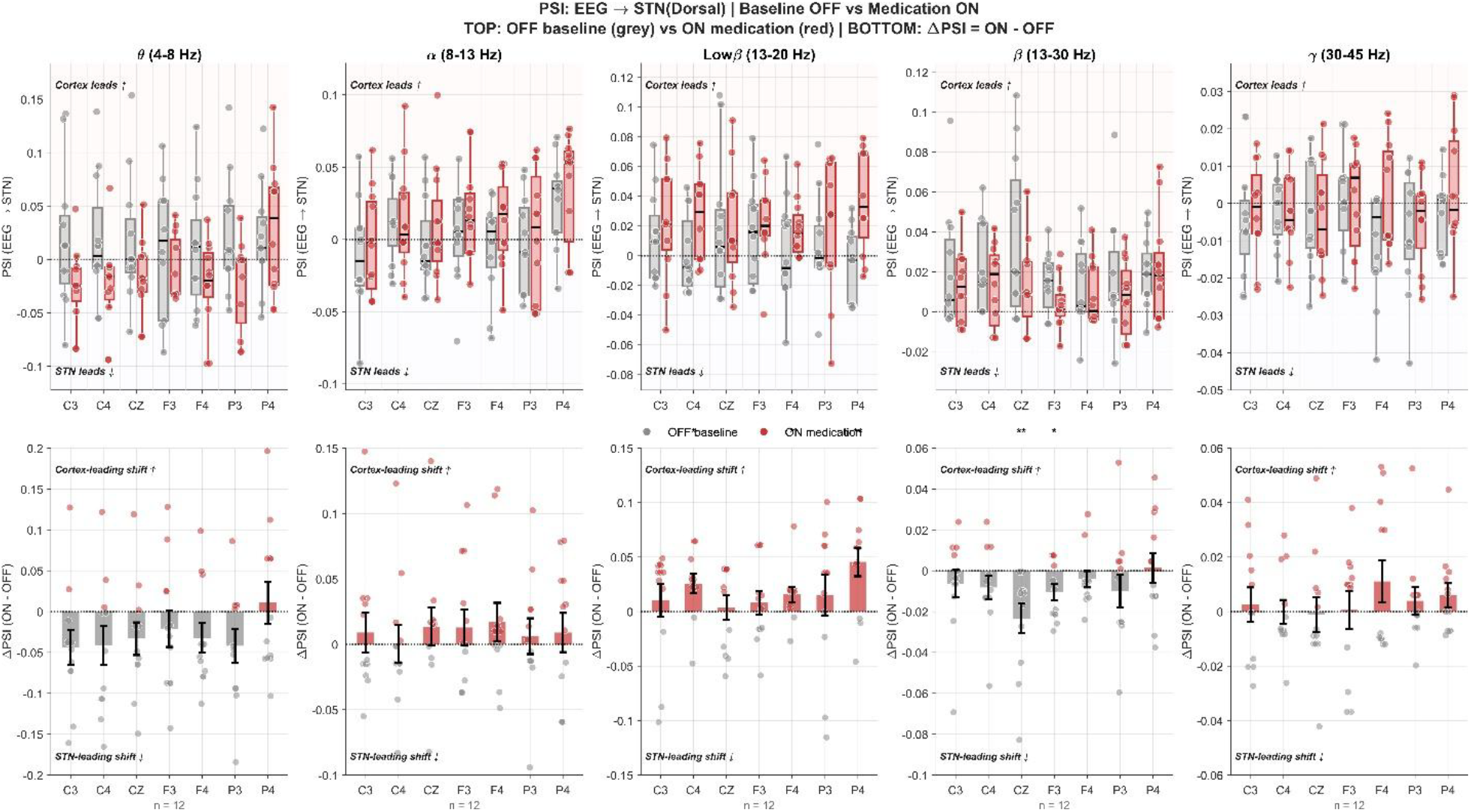
Dorsal STN: Cortical–subcortical interactions reveal cortex-driven reorganization of basal ganglia dynamics during medication. A) Top panel : Phase Slope Index (PSI) between cortical EEG electrodes and subthalamic nucleus (STN) LFPs, shown as music-induced change in directionality (Δ*PSI* = *PSI*_*Medication*_ − *PSI*_*Baseline*_). across frequency bands and recording sites. Positive values indicate cortex → STN influence; negative values indicate STN → cortex influence. B) Bottom panel: Positive ΔPSI values (warm colors) indicate cortex → STN drive, whereas negative values (cool colors) indicate STN → cortex influence.

Levodopa produced a frequency- and site-dependent alteration in directional coupling. The Δ*PSI* plots (ON − OFF; Figure 4, bottom row) showed a general shift toward positive values in the low-beta range, particularly at C4, F4, and P4. Statistically significant medication-related changes were observed at C4, F4, and P4 in the low-beta range, with the largest effect apparent at P4 (p<0.01, as indicated in the figure) and significant effects at C4 and F4 (p<0.05). These positive changes indicate that medication shifted the cortico-STN relationship toward a more cortex-leading configuration at these recording sites.

A similar pattern was observed in the broader beta range. Medication-related changes in Δ*PSI* were significant at C4 (p<0.01) and F3 (p<0.05). Thus, in the dorsal STN, the principal medication-related effect was a positive shift in cortico-STN directional coupling within the beta frequency range, suggesting increased relative cortical leading of STN activity following medication.

In theta, alpha, and gamma bands, medication-related changes were comparatively smaller and more variable across cortical electrodes, with no similarly consistent pattern of statistically significant changes apparent in the group-level analysis.

#### Ventral STN

The ventral STN recordings showed a different overall pattern from the dorsal STN (**Supplementary Figure 1)**. The baseline and levodopa-on PSI distributions were again highly heterogeneous across subjects, but the group-level PSI values were generally closer to zero in several frequency bands, indicating weaker or less consistent directional asymmetry between cortical EEG and ventral STN activity.

In the low-beta and beta ranges, medication produced relatively modest changes in Δ*PSI*, with positive and negative shifts distributed across cortical electrodes. The magnitude and consistency of these changes were smaller than those observed in the dorsal STN. Similarly, theta and alpha activity showed considerable subject-to-subject variability without a strong uniform medication-related directional shift. The gamma-band analysis also demonstrated substantial inter-individual variability, although several cortical sites had positive Δ*PSI* values following levodopa. Overall, the ventral STN analysis did not show the same robust and spatially distributed positive shift in cortico-STN directionality that was evident in the dorsal STN.

## Discussion

The principal finding of this study is that levodopa produces a spatially selective reorganization of STN physiology in PD. Rather than acting uniformly across the nucleus, levodopa preferentially modulated dorsal STN, suppressing beta power, increasing theta power, and producing larger theta–beta and gamma–beta state-space shifts, whereas ventral changes were minimal. These findings indicate that the electrophysiological response to dopamine replacement is concentrated in dorsal sensorimotor STN and is better understood as a coordinated transition between spectral states than as beta suppression alone.

PD is associated with disordered basal ganglia population dynamics, including burst firing, excessive synchronization, and persistent beta oscillations that may impair flexible motor-state transitions. Animal models provide a mechanistic foundation for this framework. In 6-hydroxydopamine-lesioned rodents, dopamine depletion alters STN firing rate and regularity, whereas chronic levodopa can normalize these firing properties; dopamine may also act directly on STN neurons through D2-like receptors^18^. In hemi-parkinsonian primates, levodopa modifies basal ganglia LFP and downstream substantia nigra pars reticulata activity even when mean STN firing rate is relatively unchanged, suggesting that therapeutic benefit depends more on firing pattern and population synchronization than on average discharge rate alone^2^. Similarly, animal studies demonstrate that acute levodopa can reverse abnormal low-frequency activity and synchronization across interconnected basal ganglia structures^20^. Collectively, these observations indicate that dopamine replacement reorganizes distributed basal ganglia dynamics rather than simply increasing or decreasing neuronal firing.

Our finding that levodopa reduced STN beta power is consistent with extensive human electrophysiological work showing excessive STN beta activity, prolonged beta bursts, and abnormal synchronization that are reduced by levodopa in proportion to improvement in bradykinesia and rigidity^22, 23, 33^. These data support the view that levodopa improves movement by destabilizing persistent beta states that oppose flexible motor updating. The present results extend this literature by showing that levodopa-related beta suppression is predominantly dorsal. This localization is anatomically and physiologically coherent with the sensorimotor dorsolateral STN, where movement-responsive neurons and beta activity are concentrated, whereas ventral associative–limbic STN generally shows lower beta and relatively greater gamma^21, 26^. High-resolution spectral mapping has further demonstrated that STN frequency biomarkers are spatially heterogeneous, with particularly strong segregation along the superior–inferior axis and a relatively superior beta hotspot^28^.

Because the pathological beta substrate is concentrated within dorsal sensorimotor STN, the stronger effect of levodopa in this region is unlikely to reflect a nonspecific nucleus-wide attenuation of activity. Instead, it suggests preferential engagement of dopamine-sensitive motor circuitry, consistent with evidence linking dorsal beta-oscillatory neurons to dopaminergic responsiveness rather than baseline impairment^21^. The present findings therefore connect two previously recognized observations—the dorsal concentration of pathological beta activity and the dopamine sensitivity of beta synchronization—by showing directly that levodopa-induced beta suppression follows STN functional topography. At the same time, the minimal ventral response should not be interpreted as evidence that ventral STN is electrically inactive or entirely dopamine insensitive. Rather, it suggests that the resting oscillatory markers examined here are substantially less dopamine responsive in ventral associative–limbic territories than in dorsal motor STN, consistent with prior evidence that beta magnitude and physiological relevance are not spatially uniform across the dorsoventral axis^25^.

Levodopa also increased theta power, again predominantly within dorsal STN. This finding is important because dopamine-related improvement is often framed largely as suppression of beta activity. However, levodopa does not simply abolish a pathological rhythm; it changes the balance among multiple competing spectral states. Low-frequency STN power has previously been shown to increase after levodopa, with the magnitude of the increase related to clinical improvement^34^. Time-resolved analyses have likewise demonstrated that levodopa reduces the occurrence and duration of low-beta states while increasing theta, alpha, and other non-beta states. A multistate model incorporating these complementary changes explains substantially more variance in motor impairment and levodopa response than beta activity alone^24^. The reciprocal dorsal reduction in beta and increase in theta observed here may therefore represent complementary components of a single therapeutic transition from a beta-dominant, relatively inflexible network state toward lower-frequency dynamics associated with state transitions and cortical–subcortical communication. Under this interpretation, theta enhancement is not merely a secondary consequence of beta suppression, but a positive feature of the reorganized neural state.

At the same time, theta activity is unlikely to have a single invariant physiological meaning. Dopaminergic therapy can modulate delta–theta activity in motor, cognitive, and affective networks ^35^. Its significance therefore depends on anatomical and clinical context. In the present study, its preferential localization to dorsal STN favors a motor-network interpretation, although concurrent behavioral and kinematic measurements will be required to determine whether dorsal theta enhancement predicts therapeutic benefit independently of beta suppression.

The theta–beta state-space analysis provides a more integrated representation of these medication effects. Levodopa produced greater dorsal than ventral displacement, reflecting a coordinated shift from high-beta/low-theta toward lower-beta/higher-theta states. This is conceptually important because neural activity is inherently multidimensional: similar single-band changes can mask substantially different overall state transitions when the coordinated behavior of other frequencies is considered. The greater dorsal theta–beta displacement therefore provides stronger evidence of a regionally specific therapeutic transition than beta or theta considered separately. It also aligns with evidence that levodopa reduces prolonged beta states and shifts multiband dynamics that predict clinical improvement better than beta power alone^23, 24^. Our results add an anatomical dimension to this framework by showing that this spectral redistribution is strongest in dorsal sensorimotor STN.

The gamma–beta analysis reinforces this interpretation. Although the change in gamma power alone did not differ significantly between dorsal and ventral STN, the combined gamma–beta projection showed a significantly greater levodopa-induced dorsal shift. This is not contradictory; a state-space projection captures coordinated movement along a composite axis, such that a significant regional difference can emerge when modest gamma changes occur together with robust beta suppression. The result indicates that dorsal STN moved farther from the dopamine-depleted configuration when beta and gamma dynamics were considered jointly. Gamma activity has frequently been associated with the dopaminergic ON state, voluntary movement, and prokinetic basal ganglia processing. Early human recordings showed that levodopa reduced low-frequency pallido-subthalamic synchronization and promoted activity near 70 Hz^10^. More recent studies have identified finely tuned gamma activity as a marker of therapeutic state and motor improvement, although its expression depends on medication, stimulation, behavior, and the presence of dyskinesia^36^. Our findings suggest that gamma may contribute to the representation of levodopa-induced network recovery even when its regional effect is insufficiently strong or consistent to serve as an independent topographic marker. In this dataset, gamma was most informative when interpreted relative to the accompanying reduction in beta activity.

Several mechanisms could account for the dorsal predominance of the levodopa response. Dorsal STN is preferentially connected with motor cortical, premotor, and basal ganglia sensorimotor territories, so dopamine restoration within striatal and related basal ganglia circuits may exert its largest downstream effect on the STN region receiving motor-loop input. Abnormal beta synchronization is also concentrated dorsally, creating a larger pathological substrate for levodopa to modulate. In addition, dopamine can influence both intrinsic STN excitability and the strength and temporal organization of afferent inputs. The observed dorsal effect may therefore reflect the combined restoration of striato-pallido-subthalamic signaling, cortical hyperdirect input, and local STN dynamics rather than a direct pharmacological effect on STN neurons alone. Prior evidence that levodopa alters functional connectivity and ventral-to-dorsal information flow within the STN supports the possibility that medication reorganizes intranuclear and network-level interactions in a regionally specific manner^27^. Dopaminergic therapy can also shift beta activity from an STN-centered motor network toward a more distributed cortical configuration^35^. Accordingly, dorsal beta suppression and theta enhancement may reflect not only local changes within STN but also altered recruitment of cortical–subthalamic and related afferent networks. The LFP changes observed here should therefore be interpreted as markers of spatially organized network reconfiguration expressed locally within dorsal STN.

These findings have translational implications for physiological localization and adaptive DBS. Beta power alone may be insufficient; dopamine-responsive dorsal STN may be better identified by combined beta suppression, theta enhancement, and multiband state-space displacement. Multiband metrics may therefore track therapeutic state more faithfully than beta alone, with biomarker thresholds calibrated to contact location. The dorsal STN region associated with optimal DBS benefit^26^ was also most levodopa responsive, suggesting a shared motor substrate that may guide contact selection and biomarker design.

Limitations include uncertainty about whether LFP effects are local or network-mediated and limited spatial resolution from overlapping DBS contact volumes. Spectral power also captures only part of STN physiology; bursts, connectivity, cross-frequency interactions, and aperiodic activity may provide additional topographic information^37^. Finally, theta/gamma are state dependent, and dorsal state-space shifts require validation against clinical improvement.

In summary, levodopa induces a spatially organized reconfiguration of STN physiology in PD. Its effects are concentrated in dorsal sensorimotor STN, where beta suppression and theta enhancement jointly produce a pronounced transition away from the dopamine-depleted neural state. The greater dorsal displacement in both theta–beta and gamma–beta state spaces shows that this transition cannot be fully characterized by any single frequency band. Together, these findings establish a topographic electrophysiological signature of dopamine replacement and suggest that levodopa restores motor function by preferentially reorganizing multidimensional network dynamics within dorsal STN.

## Data Availability

All data produced in the present study are available upon reasonable request to the authors

## Author Contributions

AShaikh, SNair: conceptualized study, collected data, analyzed data, wrote manuscript, edited manuscript.

PJoshi, ASedov, VFilyushkina, KSayfulina, AGuha, ASedov: collected data, analyzed data, edited manuscript.

## Acknowledgements and Funding

The research was supported by grants from American Parkinson’s Disease Association Cotzias Fellowship (AS), philanthropic support – Penni and Stephen Weinberg Chair in Brain Health (AS), and Dystonia Medical Research Foundation (DMRF). Shaikh was also supported by United States of Veterans Affairs RRD Merit Review (I01RX003676). The work was also supported by Russian Science Foundation (grants 23-15-00487-P and 22-15-00344-P)

## Legends

**Supplementary Figure 1.**
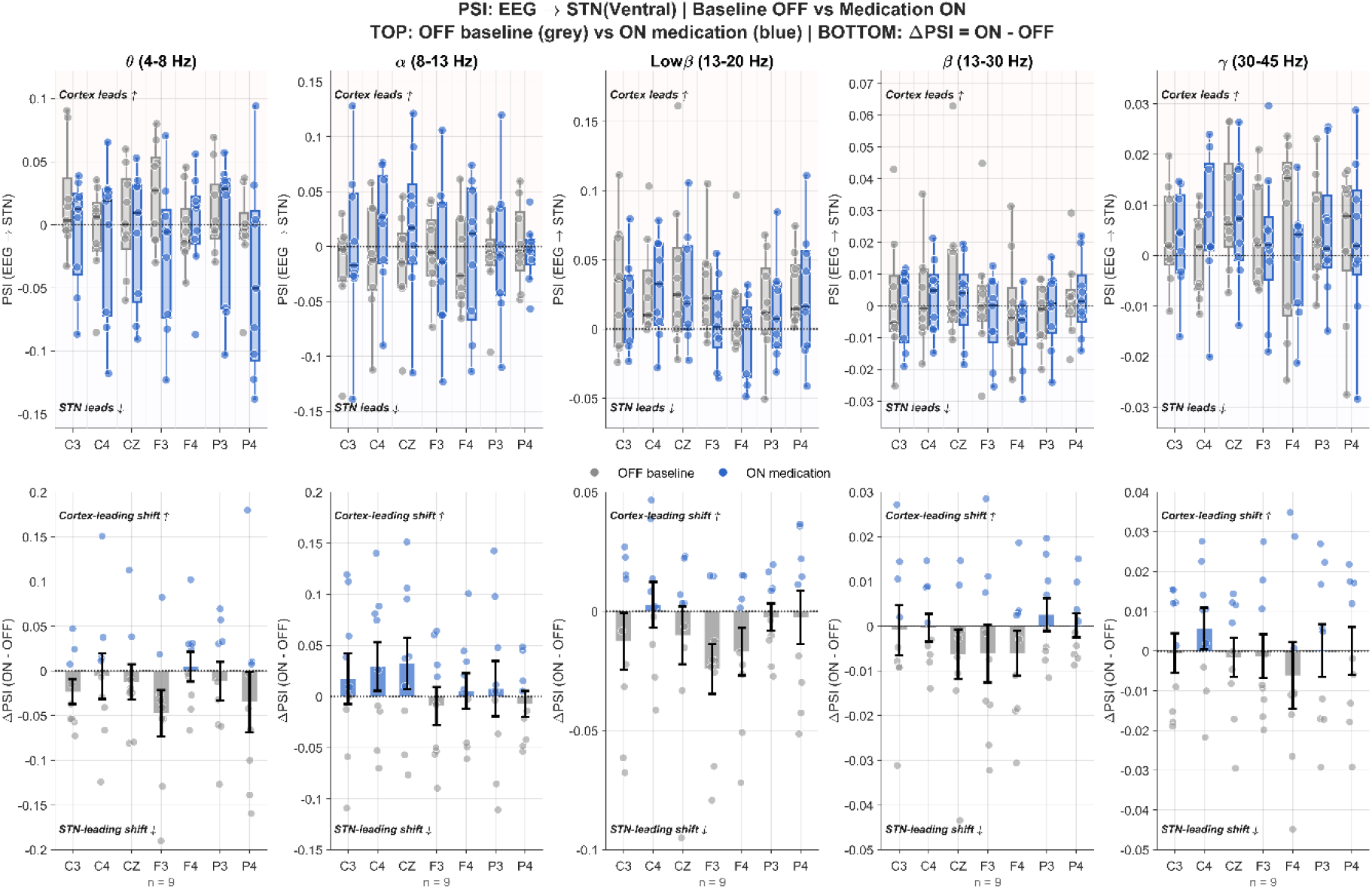
Ventral STN: Cortical–subcortical interactions reveal cortex-driven reorganization of basal ganglia dynamics during medication. A) Top panel: Phase Slope Index (PSI) between cortical EEG electrodes and subthalamic nucleus (STN) LFPs, shown as music-induced change in directionality (Δ*PSI* = *PSI*_*Medication*_ − *PSI*_*Baseline*_). across frequency bands and recording sites. Positive values indicate cortex → STN influence; negative values indicate STN → cortex influence. B) Bottom panel: Positive ΔPSI values (warm colors) indicate cortex → STN drive, whereas negative values (cool colors) indicate STN → cortex influence.

**Supplementary Table 1.** Medication-related changes in spectral power across the dorsal and ventral subregions of the subthalamic nucleus (STN). Changes in theta (ΔΘ), beta (Δ*β*), gamma (Δ*γ*), theta–beta (ΔΘ − Δ*β*), and gamma–beta (Δ*γ* − Δ*β*) power are shown for the dorsal and ventral STN, along with the corresponding p-values for comparisons between regions. Values are expressed in dB.

| Measure | Dorsal | Ventral | p-value |
| --- | --- | --- | --- |
| $\Delta\theta$ | +3.43 dB | +1.49 dB | 0.14 |
| $\Delta\beta$ | -2.47 dB | -0.12 dB | 0.0438 |
| $\Delta(\theta-\beta)$ | +5.90 dB | +1.61 dB | 0.00647 |
| $\Delta\gamma$ | +0.04 dB | +0.27 dB | 0.547 |
| $\Delta(\gamma-\beta)$ | +2.52 dB | +0.38 dB | 0.0188 |

